# Severe mental illness and last year of life: identifying service use from a national health service digital dashboard in Wales, UK

**DOI:** 10.1101/2024.11.15.24313633

**Authors:** Michael Coffey, Fiona Lugg-Widger, Ben Hannigan, Viktoriya Velikova, Anthony Byrne

## Abstract

**Objectives:** To analyse service use and variation for people with severe mental illness in the last year of life in Wales.

**Methods:** This is an observational retrospective cohort study between 2018 – 2023 using anonymised linked routinely collected health datasets within a data dashboard.

**Results:** We identified n=4722 (2.3%) deaths with ICD-10 codes for severe mental illness for the period 2018-2023. As a group people with severe mental illness die younger, are in receipt of specialist palliative care at lower rates, die more often in institutional settings rather than their own homes and comorbidity indicates more unscheduled care use in the last year of life.

**Conclusions:** Unscheduled care use in the last year of life is associated with comorbidity indicating opportunities for upstream intervention to improve treatment, experience and quality of life for people with severe mental illness. Further investigation such as mixed methodological approaches to examine experiences of those with severe mental illness in the last year of life, and the human and systems factors influencing the nature and effectiveness of unscheduled delivery for this patient cohort alongside developments in data linkage that include general practice, social care, nursing and specialist palliative care inputs are needed.

*What is already known on this topic:* There is little current data on people with severe mental illness in the last year of life, including service encounters such as unscheduled care. Routinely collected data for health service dashboards are a ready source that can provide new information on service use for this group.

*What this study adds:* We now know how many people are dying in Wales with severe mental illness and their use of services in the last year of life. Comorbidity is associated with unscheduled service use and lower rates of specialist palliative care suggest a possible upstream intervention is possible.

*How this study might affect research, practice or policy:* Detailed data linkage studies that make use of wider datasets including general practice, social care and nursing services are needed. Qualitative first-hand accounts from patients, their families and professionals could add new evidence. Specialist palliative care intervention earlier for patients with severe comorbidity may reduce use of unscheduled care.

**Funding:** The authors have not declared a specific grant for this research from any funding agency in the public, commercial or not-for-profit sectors.

**Competing interest:** VV is employed by the Digital Health and Care Wales. The Centre for Trials Research at Cardiff University receives infrastructure funding from Health & Care Research Wales. The authors declare no other competing interests.

**Data availability statement:** The dashboard has been developed by Digital Health and Care Wales in collaboration with the Welsh Value in Health Centre and NHS Wales Palliative Care clinicians. Use of the data and access to the dashboard is available to all NHS Wales staff and a number of Welsh Government professionals. All queries and requests for further information can be directed to the Information Services team at.

**Contributors:** All authors conceived and designed the study. VV prepared the dashboard data for analysis. MC performed the analysis for this paper. All authors contributed to the interpretation of the results. MC wrote the initial draft of the manuscript and all authors critically reviewed and approved the final version.

## Introduction

People with severe mental illness (SMI) diagnoses such as schizophrenia or bipolar disorder are known to have a life expectancy some 20 years shorter than the general population [1]. For those with an additional life limiting illness such as cancers or organ failure, there is a complex set of inequalities that lead to a mismatch between their care needs and service provision [2]. Variation in care is attracting international research efforts to better understand care use and outcomes [3,4]. For people with severe mental illnesses multiple challenges exist in accessing better end-of-life care with little research directly estimating the size of the cohort, service use, and outcomes suggesting probable significant unscheduled care use [5].

Routinely collected data produced in healthcare settings present an important opportunity for addressing research questions and to inform planning in health services [6]. These data can address knowledge gaps, generate hypotheses [7], help in developing new tools for promoting quality and safety in healthcare [8], contribute to assessing impact of healthcare interventions [9] and allow evaluation of effectiveness to inform policy decisions [6]. Digital dashboards are increasingly being used as a means of data visualization in healthcare systems to communicate knowledge of utilization and prompt data-based service improvement [10]. Where these dashboards are easily accessible to clinicians their use is associated with improved patient care and better outcomes [11]. This paper examines data arising from one such dashboard that collates last year of life data at a population level for the country of Wales, UK. Our research question is, ‘What can a national data dashboard tell us about the use of services in the last year of life for the population of people with severe mental illness in Wales?’

Systematic reviews indicate that healthcare systems contribute to numerous challenges for those with severe mental illnesses in negotiating access and securing coordinated care [5], that service use in the context of co-morbidity varied but was likely to be higher for this group [12] and more recently that co-morbid cancer diagnoses may lead to increased use of palliative care but less high intensity end-of-life care [4]. An earlier review did not identify any comparable UK data on incidence, prevalence, or service use for people with SMI diagnoses to inform supportive care responses [5]. Nevertheless, health services are routinely collecting and making use of data that can provide useful insights into what is happening for specific patient groups. Digital dashboards capture, analyse and present data on how a given service is performing to allow optimisation of services and enhance their performance [13].

The last year of life digital dashboard used in this paper is provided by the Wales Value in Health Centre and Digital Health and Care Wales. Value in Health is a National Health Service (NHS) Wales programme focused on delivering value-based healthcare across healthcare delivery with a focus on outcomes that matter to patients and a more data-driven system. Digital Health and Care Wales is part of NHS Wales and uses data to improve how health and care services are delivered.

Our objectives were to use the last year of life dashboard to describe this population, address the research knowledge gap of service use in the last year of life for people with severe mental illness in a whole population, and finally, to understand unscheduled care use in this cohort. We also wanted to learn what additional data would be needed to build a more complete picture of last year of life for this group.

## METHODS

### Study Design

This is an observational retrospective cohort study using multiple routinely collected health datasets within a data dashboard. This study is reported using the RECORD reporting guidelines [14]. An eligible ICD-10 cohort was produced (VV) in the last year of life dashboard and information was aggregated to years 2018-2021 and 2022-2023. We examined data relating to deceased people for the whole of Wales and a sub-population of patients with SMI diagnoses.

#### Population

All recorded deaths for the database population (defined as everyone registered with a Welsh general practice and/or everyone identified as living in Wales at the time of death from Office of National Statistics data) in Wales for the years 2018-2023, and from these data a cohort (study population) of all deceased people with a record of severe mental illness, identified as any lifetime diagnosis using ICD-10 codes for schizophrenia, schizotypal and delusional disorders F20-29, mood disorders F30-39, disorders of adult personality and behaviour F60-69 [15]. Deaths due to unnatural causes, e.g. accidents were excluded as these patients would not typically be candidates for palliative care. Linkage between datasets used individual NHS numbers and the time frame was set to 12 months prior to the patients’ deaths.

#### Data Sources

The following anonymised data sets were used to generate linked data outputs on the dashboard.

- NHS 111 calls data set – this is a National Health Service urgent call line for advice and if needed onward referral to emergency department, urgent treatment centre or pharmacy.
- General Practice (GP) out of hours data set – calls to general practice out of hours.
- Welsh Ambulance Service data set – calls and ambulances dispatched.
- Welsh Ambulance Service patient clinical record (PCR) data set - PCR data from ambulance crews (data from 2018-2021).
- Emergency department data set – emergency departments and minor injuries unit data in Wales.
- Admitted patient care data set –consultant-led admitted patient care data (elective and emergency).
- Office of National Statistics Death data set – monthly death registration data.
- Welsh Demographic Service data set – data on all patients registered with GPs in Wales.
- Cancer Network Information System Cymru (CaNISC) –cancer patients and specialist palliative care (SPC) provision data in community and secondary care.

#### Permissions

Access to the dashboard created by Digital Health and Care Wales was granted in April 2024, on the basis of controls that limit the risk of personally identifiable information being available and suppression of low numbers. Research ethics approval was not applicable as this was a service evaluation. Values relating to 5 or less patients were suppressed in all dashboard visualisations as a privacy protection method.

#### Analysis

We described the cohort of people with severe mental illness diagnoses dying in Wales compared with the wider Welsh population, in particular:

- Demographic variables included age at death and sex.
- Underlying cause of death using ICD-10 diagnosis codes e.g., diseases of the circulatory system, neoplasms etc.
- A delineated count of care episodes, scheduled and unscheduled, in the last 12 months and last 90 days of life (A&E, Ambulance, Emergency bed days, and NHS 111 service use).
- The number of days spent in hospital (admission to discharge date) in the last 12 months of life.
- Place of death (e.g. home, care home, NHS establishment, other).
- Provision of specialist palliative care.
- A Charlson Comorbidity Index (CCI) score 16 was derived to enable an estimate of the associated impact of co-morbidities on unscheduled health care use for those with SMI diagnosis.

## Results

The most recent dashboard data indicates 89.3% average clinical coding completeness across all Welsh Health Boards for 2022-2024 with 180k total uncoded episodes.

### Demographics

The last year of life dashboard data identified 204,576 recorded deaths between 2018-2023 (table 1). Of these 2.3% (n=4722) had an ICD-10 code indicating a lifetime diagnosis of a severe mental illness. On average people with SMI diagnoses die younger than others in the population (SMI 37% vs non-SMI 20% in the 18-69 years group), are in receipt of specialist palliative care at lower rates, die in hospital at similar rates and have lower percentage of severe comorbidities and more ratings for mild or no comorbidities. On average the SMI cohort use emergency and ambulance services, as a cohort at similar rates as the non-SMI group but use of emergency beds is higher.

**Table 1:** Showing demographic information for last year of life in the cohort of people with SMI diagnoses and the whole population.

|  | *Non-SMI*<br>*n=* | *%* | *SMI*<br>*n=* | *%* | *Total* | *%* |
| --- | --- | --- | --- | --- | --- | --- |
| *Deaths* | 199,854 | 97.6 | 4,722 | 2.3 | 204,576 | 100 |
| *Age groups* |  |  |  |  |  |  |
| *<18 yrs.* | 942 | .4 | 0 | 0 | 942 | 1 |
| *18-69 yrs.* | 39,851 | 19.9 | 1,742 | 36.9 | 41,593 | 20 |
| *70 +* | 159,061 | 79.5 | 2,980 | 63.1 | 162,041 | 79 |
| *Sex* |  |  |  |  |  |  |
| *Male* | 99,853 | 49.9 | 2,155 | 45.6 | 102,008 | 49.9 |
| *Female* | 100,001 | 50 | 2,567 | 54.3 | 102,568 | 50.1 |
| *Specialist Palliative Care* | 66,964 | 33.5 | 1,169 | 25 | 68,133 | 33.3 |
| *Dying in hospital* | 101,238 | 50.6 | 2,271 | 48.1 | 103,509 | 50.5 |
| *5-year Charlson comorbidity index\** |  |  |  |  |  |  |
| *Severe* | 66,004 | 33 | 1,234 | 26 | 67,238 | 33 |
| *Moderate* | 45,412 | 22.7 | 1,058 | 22 | 46,570 | 28 |
| *Mild* | 50,554 | 25.2 | 1,522 | 32 | 52,072 | 23 |
| *None* | 32,788 | 16.4 | 908 | 19 | 33,696 | 16 |
| *Emergency department contacts in last 365 days* | 292,234 | 43.6 | 8,956 | 41.8 | 301,190 | 43.5 |
| *Ambulance call outs Last 365 days* | 263,682 | 39.3 | 8,794 | 41.1 | 272,476 | 39.4 |
| *Emergency Bed admissions last 365 days* | 295,811 | 53.4 | 7,813 | 74 | 303,624 | 53.8 |

\*Charlson Comorbidity Index groups by score: 0=no CCI, 1-2=mild, 3-4=moderate, >=5 = severe.

Underlying cause of death: The top three causes of death by ICD-10 Chapter for the SMI group were diseases of the circulatory system n=1145 (24%), diseases of the respiratory system n=837 (17.7%), and neoplasms n=834 (17.6%). This compares with the non-SMI group where neoplasms n=56,293 (27.5%), diseases of the circulatory system n=51,769 (25%), and diseases of the respiratory system n=27,333 (13%).

### Care episodes

People with SMI had a total of 33,885 care encounters in the last year of life, with 14,236 (44%) of these contacts occurring in the last 90 days of life. The majority of contacts for the SMI group are distributed between ambulance call outs and emergency department attendance along with admitted patient care episodes (Table 2). Primary care out of hours encounters are higher in the SMI group in the last year of life. Compared with the non-SMI group the SMI group show a higher percentage of total care encounters with unscheduled care and fewer attendances of less than one day in the last year of life. Encounters on day of death for both groups are similar (Table 3). Specialist palliative care contacts indicate variance between SMI and non-SMI groups with a lower percentage of contact for the SMI grouping (SMI n=1932, 5.7% versus non-SMI n=113,829, 8.5% in last year of life). These are not included in table 2 to avoid confusion as the dashboard data only indicates initial contact and not ongoing or repeated contacts for specialist palliative care.

**Table 2:**
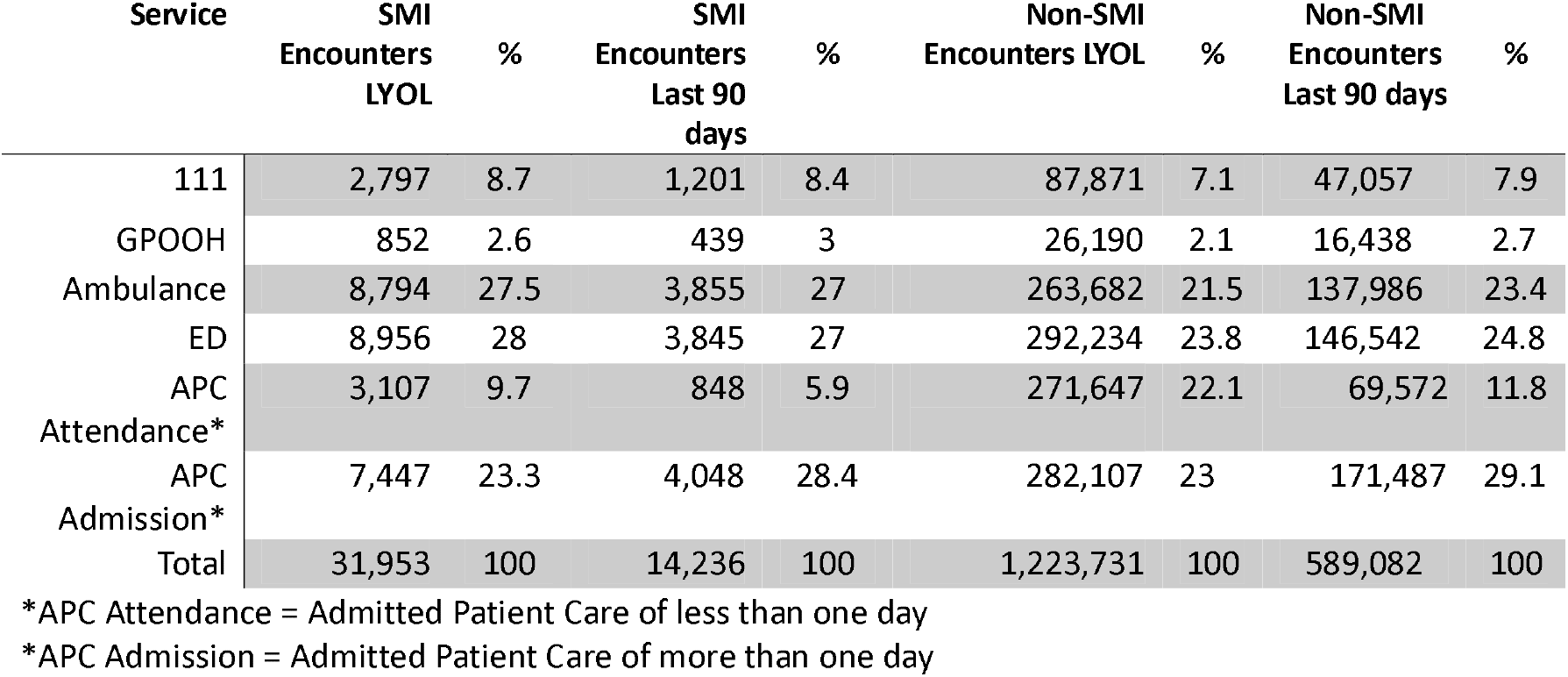
SMI vs Non-SMI care encounters in last year of life and last 90 days of life.

**Table 3:**
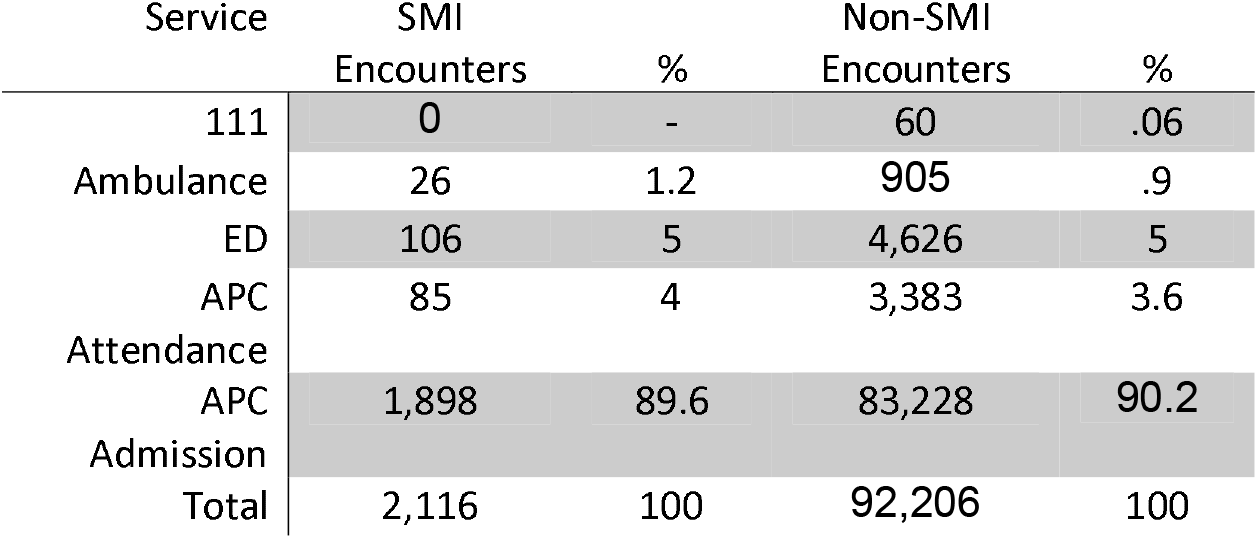
SMI versus non-SMI contacts on day of death.

### Days spent in hospital

Emergency bed days are higher on average for SMI compared with the non-SMI population in the last year of life (see table 1) and this difference remains for the last 90 days with the SMI group mean for emergency bed days higher in comparison (23 vs 19 bed days).

### Place of death

Most people in both groups die in formal settings such as a hospital or care home (see Table 4) with the SMI group dying showing a higher percentage of dying in a care home and lower percentage for dying in their own home than the non-SMI group. Hospice as place of death was lower on average for the SMI group.

**Table 4:** SMI and Non-SMI place of death.

| Place of death | SMI | % | Non-SMI | % |
| --- | --- | --- | --- | --- |
| Hospital | 2,271 | 48 | 101,244 | 50.6 |
| Care Home | 1,138 | 24.1 | 33,953 | 16.9 |
| Home | 1,095 | 23.1 | 56,067 | 28 |
| Hospice | 66 | 1.4 | 4,441 | 2.2 |
| Other | 152 | 3.2 | 4,149 | 2 |
| Total | 4722 | 100 | 199,854 | 100 |

### Specialist palliative care

The total number in receipt of SPC in the SMI cohort was n=1169 (25%) and their earliest encounter with SPC was on average 98 days prior to death with a median of 35 days. Compared with non-SMI group the SMI group had less specialist palliative care on average in the last year of life. For 18-69 years receipt of SPC in the non-SMI group was 41% versus 26.8% for the SMI group and for 70 years and above the non-SMI group was 31.4% versus 23.5% for the SMI group.

In the SMI group just under half (48%, n=2271) died in hospital, of these 73% (n=1660) had no SPC versus 26% (n=611) who had received SPC indicating an association between SPC receipt and dying in hospital. The cohort in receipt of SPC had a higher percentage of cause of death recorded as neoplasm (n=604), usually metastatic in nature. In the SMI cohort the total number of bed days (see Figure 1) for those with SPC was less than half (47.7%) that of those with no SPC (52,492 days vs 110,138 days).

**Figure 1.**
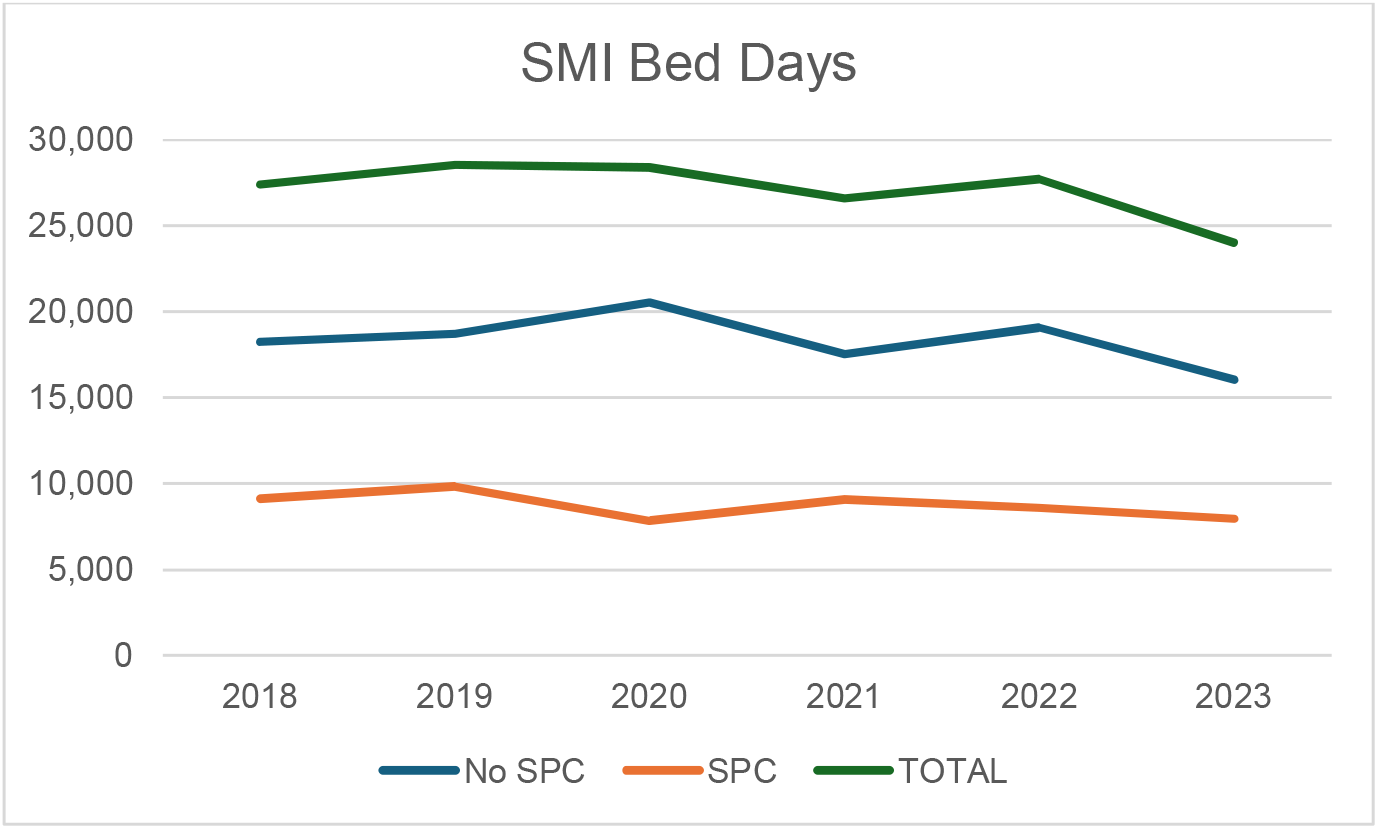
Showing bed day count for the SMI cohort for those with and without specialist palliative care.

### Co-morbidities

Table 1 reports the 5-year CCI scores for the SMI cohort versus the whole population. Unscheduled service encounters in the last year of life for those with SMI and CCI scores of 5 or more (severe) are 7,600 contacts for n=1,234 deaths. Those with more comorbidities have most unscheduled service encounters e.g., those with no comorbidities (n=908) accounted for only 8% (n=1826) of unscheduled encounters versus 35% (n=7600) for those with most comorbidities in the last year of life.

Proportionately more contacts occur in the last 90 days of life, e.g. 41% of all unscheduled encounters for those with a CCI severe score occur in the last 90 days with most encounters with ambulance and emergency departments (Table 5). Unscheduled care encounters are in the expected direction, i.e., higher CCI scores are in line with those with most encounters, with the exception of those with CCI mild (scores of 1-2) with the largest number of deaths and the second highest number of unscheduled care encounters.

**Table 5.** Unscheduled care encounters by CCI score in last year and last 90 days of life for the SMI cohort.

|  | *CCI Severe<br>Last 365: Last 90<br>days<br>N=1,234 | CCI Moderate<br>Last 365: Last 90<br>days<br>N=1,058 | CCI Mild<br>Last 365: Last 90<br>days<br>N=1,522 | CCI None<br>Last 365: Last 90<br>days<br>N=908 |
| --- | --- | --- | --- | --- |
| *All encounters | 13,536: 5,858 | 8,011: 3,874 | 9,794: 4,670 | 2,544: 1,248 |
| WAST | 3,034: 1,246 | 2,320: 1,008 | 2,744: 1,248 | 696: 353 |
| ED | 3,151: 1,298 | 2,190: 966 | 2,848: 1,237 | 767: 344 |
| NHS 111 | 1,086: 436 | 617: 282 | 800: 363 | 294: 120 |
| GPOOH | 329: 154 | 185: 111 | 269: 126 | 69: 48 |

\*Charlson Comorbidity Index groups by score: 0=no CCI, 1-2=mild, 3-4=moderate, >=5 = severe.
\*All contacts total includes contacts such as admissions where it is not clear if these were elective or emergency.

## Discussion

We explored service use in the last year of life for people with a lifetime diagnosis of SMI using the last year of life dashboard for Wales, UK. For the first time we have been able to identify a whole country SMI cohort (n=4,722) over a six-year period and follow their service use in the last year of life across a number of acute services. Our results show that this group die younger than the non-SMI group, have more unscheduled care episodes, die more often in either hospital or in a care home (72% vs 67.5%), spend more time in hospital in general and are in receipt of specialist palliative care at lower rates. In the SMI cohort, those that received specialist palliative care had substantially less hospital bed days indicating a possible effect for this care. Additionally, we can observe that those with comorbidities make more use of unscheduled care services (88.5% vs 11.5%), and that in the last 90 days of life those with comorbidities account for 87% of all unscheduled contacts for this cohort. These results are similar to those found in other countries. In a Swedish study those with psychosis and cancer were less likely to receive palliative care than those without psychosis, died on average 4 and half years younger and when specialist palliative care was provided this led to reduced frequency of emergency care use [17]. Comorbidities are also implicated with an associated increased use of unscheduled care in a London cohort [18].

The experience of severe mental illness can delay detection and treatment of physical disorders as individuals are less likely to seek treatment, verbalise pain and access timely healthcare [19]. Decreased engagement with medical care in comparison with the general population can be due to psychological and social issues, poor previous experiences of seeking help, healthcare professionals wrongly attributing physical symptoms to psychiatric disorders and lack of experience by mental health staff in determining how and when to refer to appropriate services [20]. In this study we have identified significant use of unscheduled care implying ad hoc, chaotic and reactive engagement indicated absent, reduced or missed opportunities for planned medical care. People with enduring mental health problems are approximately 50% less likely to access appropriate palliative care, including symptom control and pain relief [20]. This is despite a well-recognised outcome indicating that earlier palliative care is associated with increased survival, better quality of life, and less need for costly intervention at the end of life [21,22].

Our data on people with SMI is the first indication of service use in the last year of life for this group in the UK. They are dying younger and in service settings, they are in receipt of less specialist palliative care, are using unscheduled care at higher rates and this appears in part related to presence of comorbid conditions. Together these data indicate that intervention could be targeted at the SMI group with comorbidities not just with the aim of reducing costly unscheduled care use but to provide appropriate evidenced-based care, improve quality of life and remove variation [23].

Complexity is a widely acknowledged feature of public health systems and the service provision and care experiences of people with severe mental illness is a particular example of these challenges [24, 25]. Wider social barriers arise for people with mental health problems who also have a life-limiting illness which may defy simple, linear approaches to researching cause and effect. As a group people with severe mental illness are more likely to be socially isolated and to be homeless, which impacts upon care planning[26]. Inadequate support systems are common for this group, which affects their ability to access care and navigate complex health and social care systems, likely increasing unscheduled care [27]. These challenges play out in the ways people interact with services and the care that is provided to them. For example, the National Cancer Patient Experience Survey showed that people with severe mental illness were least likely to say they got the right support with cancer care [28]. Although incidence of cancer was similar to the general population in a Swedish study, mortality rates for those with a mental health problem and cancer were double [29]. This disparity may be related in part to late presentation and reduced use of interventions such as surgery, chemotherapy, or radiotherapy [30]. Data available in Wales indicates most people who need it do not receive specialist palliative care [31]. Of those with cancer, 46% access specialist palliative care and for other life limiting conditions, this is only 5.3%. Emergency hospital attendance is higher for those with other conditions compared to cancer (85-89% versus 65%), and three quarters of people are admitted at least once to hospital in the last year of life.

### Strengths and limitations

A strength of this study is that we were able to exploit a rich source of patient-level data for a whole country and present for the first time analysis indicating variation in service use for the SMI population that experience significant inequity in care provision. There are limitations to this evaluation arising from those that apply more broadly to data from digital dashboards. For example, dashboards may direct attention towards certain data while important areas may be neglected.

Social care and routine primary care data are not included in the dashboard to date and therefore the picture we present while important remains focused on unscheduled care with only partial access to other data. We were also unable to determine if the SMI cohort were in receipt of continuing secondary care from mental health services. Our analysis is limited too by data availability. The dashboard does not report ethnicity data or living situation, and this reduces what we can say about the social circumstances of individuals. Dashboards are mainly visualisation tools. Our permissions did not include access to underlying data, and this therefore limits our ability to manipulate data for inferential purposes. Routinely collected data imported into a secure environment would allow more sophisticated analyses including generating a matched cohort for comparisons. We recognise too that the last year of life dashboard does not distinguish between types of hospital admissions limiting our ability to further determine variation. Generalisability beyond the UK is limited to similar healthcare systems. Lifetime diagnosis was gained from secondary care recording of illness, however those never seen in secondary care, or whose SMI were not captured on death certification, will not have been identified and hence we consider to be underestimates.

## Conclusion

The current paper is a first step in understanding how people in Wales with severe mental illness interact with the health system and the variation that occurs when they do. Further research is required to address questions about the how and why of their interactions. These include mixed methodological approaches which give in depth understanding of lived experience of those with SMI in the last year of life, and the human and systems factors which determine the nature and effectiveness of unscheduled delivery for this patient cohort. Continued development of datasets will allow impact of new interventions to be assessed over time, with improved understanding of how the wider systems of care respond and adapt. There is need too for detailed data linkage studies that include social care and nursing care data points to fully understand the effect of supportive care on unscheduled care use. Nevertheless, these data show that the SMI cohort die younger, experience variation in care (less SPC) and make more use of unscheduled care indicating possible opportunities for intervention to remove inequalities for this group. These data underpin the utility of population level datasets and mandates for widening the types of data available for integrated analysis with a view to iterative complex service improvements.

## Acknowledgements

The authors acknowledge the support and assistance of Digital Health and Care Wales in making the dashboard available for analysis.

